# Evaluating the Association between Routine Pneumococcal Vaccination and COVID-19 Severity among Older Adults in the United States

**DOI:** 10.1101/2023.12.27.23300578

**Authors:** Ottavia Prunas, Andrew Tiu, Shweta Bansal, Daniel M. Weinberger

## Abstract

**Background:** The relationship between severe acute respiratory syndrome coronavirus 2 (SARS-CoV-2) and *Streptococcus pneumoniae* remains uncertain. This study investigates the association between routine pneumococcal vaccination and the progression to severe COVID-19 outcomes in a cohort of older adults in the United States.

**Methods:** Our cohort study includes adults aged 65 and older from a subset of adults covered by Medicare in the United States with a documented COVID-19 diagnosis. Logistic regression models were employed to assess the association between pneumococcal vaccination (13-valent conjugate vaccine [PCV13] and 23-valent pneumococcal polysaccharide vaccine [PPSV23]) and COVID-19 severity.

**Results:** Among 90,070 Medicare enrollees with a COVID-19 diagnosis, 28,124 individuals exhibited severe respiratory symptoms or were admitted to the intensive care unit (ICU). The odds ratio (OR) for progression from non-severe symptoms to respiratory symptoms with or without ICU admission with prior PCV13 receipt was 0.91 (95% confidence interval [CI], 0.88, 0.93), the OR for progression from severe respiratory symptoms to ICU critical care with prior PCV13 receipt was 0.92 (95% CI, 0.88, 0.97), and the OR for progression from non-severe symptoms to ICU critical care with prior PCV13 receipt was 0.85 (95% CI, 0.81, 0.90). There was no association between PPSV23 received more than five years before the COVID-19 diagnosis and the COVID-19 outcomes.

**Conclusions:** Overall, our findings indicate moderate to no association between PCV vaccination and COVID-19 severity.

## Introduction

*Streptococcus pneumoniae*, or pneumococcus, significantly contributes to the burden of lower respiratory tract infections (LRTIs), a substantial cause of morbidity and mortality worldwide [1, 2]. Pneumococcus interacts synergistically with respiratory viruses, particularly influenza and respiratory syncytial virus, creating an inflammatory environment that promotes pneumococcal carriage and weakens the host’s defense against secondary bacterial pneumonia [3–9].

Pneumococcal conjugate vaccines (PCVs) protect against a subset of pneumococcal serotypes by reducing upper respiratory tract colonization and the risk of severe disease. Randomized clinical trials (RCTs) and epidemiological studies suggest that PCVs may also reduce the risk of viral lower respiratory tract infections in both children and adults by preventing co-infections [6, 10–14].

Recent research has explored interactions between SARS-CoV-2 and pneumococcus [15], yielding mixed evidence. While some studies suggest a potential protective effect of 13-valent conjugate vaccine (PCV13) on COVID-19 diagnosis [6, 16], others fail to substantiate such a connection [17]. To contribute further to the topic, our study investigates potential interactions between SARS-CoV-2 and pneumococci. We achieve this by examining the severity of COVID-19 outcomes in a large cohort of older adults in the United States, differentiating between those who have received PCV13 and 23-valent pneumococcal polysaccharide vaccine (PPSV23) vaccinations and those who have not.

## Methods

### Data sources

We used Limited Data Set (LDS) files from the Centers for Medicare & Medicaid Services (CMS) for claims from inpatient facilities, outpatient facilities, skilled nursing facilities, and doctors’ offices. Combined, the claims data spanned the period from 2014 to 2021. Additionally, we had access to the age and race/ethnicity of all beneficiaries. We formed the study cohort by including patients who were continuously and completely observed from 2014 to 2021 in the available LDS data and had a first COVID-19 diagnosis (using International Classification of Diseases, ICD-10 code criterion) anytime from April 4, 2020 to December 31, 2021 (n = 90,070) (See Figure S1 and Supplementary Information for details). Because individuals had to be observed for this entire period, and most people become eligible for Medicare at age 65, individuals aged <72 were under-represented in the cohort. For the patients in the cohort, we characterized COVID-19 outcome severity based on four mutually exclusive groups of increasing severity which were defined based on diagnoses that co-occurred with COVID-19 diagnosis (on the same claim) up to six months after the initial COVID-19 diagnosis (ICD-10 criteria in Table S1): non-severe (COVID diagnosis with none of the diagnoses in the other categories), non-respiratory severe (including stroke, cardiomyopathy, myocardial infarction, heart failure, and distress); respiratory severe (including pneumonia, bronchitis, respiratory distress, and respiratory failure); and critical care (including the need for ventilator or critical care services). We also identified whether patients had received pneumococcal conjugate vaccination, influenza vaccination, and zoster vaccination (using vaccine-specific Current Procedural Terminology (CPT) or Healthcare Common Procedure Coding System (HCPCS) code-based criteria) provided that the vaccination occurred in a claim before the initial COVID-19 diagnosis (Table S2). We additionally characterized the comorbidities of each patient, and whether they experienced past severe respiratory illness (based on ICD-10 criteria, Table S3).

### Statistical analysis

We employed a series of logistic regression models to assess the association between receipt of pneumococcal vaccination (PCV13 and/or PPSV23) and severity of COVID-19. The models evaluated associations between the odds of having more severe versus less severe outcomes as a function of pneumococcal vaccine status. Based on our definitions of COVID-19 outcome severity, we evaluated the risk of progression from less severe to more severe categories through the following models:

Model 1: odds of severe respiratory outcomes or critical care versus non-severe outcomes.
Model 2: odds of severe respiratory outcomes or critical care versus severe non-respiratory outcomes.
Model 3: odds of critical care versus severe respiratory outcomes. Model 4: odds of critical care versus non-severe outcomes.

The main exposure of interest was whether the individual had received any dose of PCV13; any dose of PPSV23 within the past five years; or any dose of PPSV23 more than five years ago, without a dose of PPSV23 within the last five years. The analysis was limited to individuals aged 65 and above, grouped into six-year age intervals: 65-69, 70-74, 75-79, 80-84, 85-88, and 89+. We adjusted for race/ethnicity (Table 1) and comorbidities. We also included a covariate indicating whether the patient had a severe respiratory diagnosis code (Table S1) in the period from 2015 to the year before their initial COVID-19 diagnosis. To investigate potential variations in the associations between pneumococcal vaccination (PCV) exposure and COVID-19 severity concerning different COVID-19 variants (i.e., original strain, Alpha variant, and Delta variant), we incorporated a time period indicator variable ((1) between April 2020 and August 2020 for the original strain; (2) between September 2020 and March 2021 for the Alpha variant; and (3) between April 2021 and December 2021 for the Delta variant). As negative controls, we evaluated the associations between receipt of influenza or zoster vaccinations and the outcomes, substituting these vaccines for the pneumococcal vaccines in the model. Influenza was only circulating at very low levels during much of the study period, so we would expect no effect.

**Table 1:** Characteristics of the Study Cohort Individuals.

| No. (%) of individuals |  |  |  |  |  |  |
| --- | --- | --- | --- | --- | --- | --- |
|  | Unvaccinated | PCV13 only | PPSV23 only<br>(last 5 years) | PPSV23 only<br>(> 5 years) | PCV13+<br>PPSV23<br>(last 5 yrs) | PCV13+<br>PPSV23<br>(>5 yrs) |
|  | n=32,494 | n=27,981 | n=5,898 | n=3,039 | n=15,761 | n=4,897 |
| Age |  |  |  |  |  |  |
| 65-69 years | 2408 (7) | 802 (3) | 676 (11) | 193 (6) | 798 (5) | 135 (3) |
| 70-74 years | 6984 (21) | 4585 (16) | 1394 (24) | 786 (26) | 4340 (28) | 1662 (34) |
| 75-79 years | 8035 (25) | 7736 (28) | 1379 (23) | 786 (26) | 4295 (27) | 1280 (26) |
| 80-84 years | 6262 (19) | 6200 (22) | 1115 (19) | 520 (17) | 3080 (20) | 835 (17) |
| 85-89 years | 4459 (14) | 4670 (17) | 746 (13) | 387 (13) | 1849 (12) | 562 (11) |
| 90+ years | 4346 (13) | 3988 (14) | 588 (10) | 367 (12) | 1399 (9) | 423 (9) |
| History of respiratory symptoms before the Covid-19 diagnosis |  |  |  |  |  |  |
| No | 13307 (41) | 9962 (36) | 1885 (32) | 1065 (35) | 5281 (34) | 1676 (34) |
| Yes | 19187 (59) | 18019 (64) | 4013 (68) | 1974 (65) | 10480 (66) | 3221 (66) |
| Race/ethnicity |  |  |  |  |  |  |
| Unknown | 322 (1) | 217 (1) | 67 (1) | 34 (1) | 207 (1) | 76 (2) |
| White | 26797 (83) | 25175 (9) | 4654 (79) | 2509 (83) | 13872 (88) | 4306 (88) |
| Black | 3370 (10) | 1434 (5) | 665 (11) | 255 (8) | 973 (6) | 262 (5) |
| Other | 429 (1) | 262 (1) | 100 (2) | 36 (1) | 189 (1) | 58 (1) |
| Asian | 557 (2) | 332 (1) | 142 (2) | 87 (3) | 193 (1) | 78 (2) |
| Hispanic | 793 (2.4) | 383 (1) | 217 (4) | 84 (3) | 258 (2) | 94 (2) |
| Native American | 226 (0.7) | 178 (1) | 53 (1) | 34 (1) | 69 (0.004) | 23 (0.005) |
| Comorbid conditions |  |  |  |  |  |  |
| No | 2717 (8) | 2724 (9.7) | 332 (6) | 180 (6) | 1341 (9) | 508 (10) |
| Yes | 29777 (92) | 25257 (90) | 5566 (94) | 2859 (94) | 14420 (91) | 4389 (90) |
| Influenza vaccination |  |  |  |  |  |  |
| No | 31277 (96) | 1685 (6) | 501 (8) | 332 (11) | 640 (4) | 4683 (96) |
| Yes | 1217 (4) | 26296 (94) | 5397 (92) | 2707 (89) | 15121 (96) | 214 (4) |
| Zoster vaccination |  |  |  |  |  |  |
| No | 32470 (100) | 27130 (97) | 5761 (98) | 2977 (98) | 15228 (97) | 253 (5) |
| Yes | 24 (0) | 851 (3) | 137 (2) | 62 (2) | 533 (3) | 4644 (95) |

For the main effects, we present the point estimate for the odds ratio and the 95% confidence interval. All analyses were performed in R version 4.1.2 (Vienna, Austria). The main code can be found in the following github repository: https://github.com/weinbergerlab/PCV-analysis.

## Results

The study cohort consisted of 90,070 individuals aged 65 years and older enrolled in Medicare health plans who had a COVID-19 diagnosis and were observed from 2014 to 2021 (Figure S1). Of these, 8,067 individuals were admitted to ICU critical care, 20,057 exhibited severe COVID-19 respiratory symptoms, 7,336 individuals experienced severe non-respiratory COVID-19 outcomes, and 54,610 had a diagnosis of COVID-19 without outcomes in the other severe classes. In total, 48,639 (54%) individuals received PCV13 with or without PPSV23 at the age of 65 or older. Of these, 27,981 individuals received PCV13 only, 15,761 individuals received PPSV23 within the five years before their COVID-19 diagnosis, and 4,897 also received PPSV23 more than five years before their COVID-19 diagnosis. 8,937 (9.9 %) individuals received PPSV23 only; of these, 5898 individuals received PPSV23 within five years before their COVID-19 diagnosis, while 3,039 received PPSV23 more than five years before their COVID-19 diagnosis. Notably, individuals who received any pneumococcal vaccine were generally older than non-recipients and exhibited a higher prevalence of comorbid conditions associated with pneumonia and COVID-19 risk, along with pre-existing respiratory symptoms before the COVID-19 diagnosis (Table 1).

Overall, there was moderate association between the receipt of pneumococcal vaccines and COVID-19 severity for some outcomes, and no association for other COVID-19 outcomes (Figure 1 and Table 2). Specifically, we identified a modest association between receipt of PCV13 only and the odds of having severe respiratory or critical care outcomes compared with the odds of having non-severe outcomes (Outcome 1) (odds ratio, OR: 0.91 with 95% Confidence Interval, CI: 0.88,0.93). Likewise, there was a modest association between PCV13 receipt and odds of critical care versus odds of severe respiratory outcomes (Outcome 3) (OR: 0.92, with 95%CI: 0.88,0.97), and between PCV13 receipt and odds of critical care versus odds of non-severe symptoms (OR: 0.85, with 95%CI: 0.81,0.90). In contrast, receiving PPSV23 more than five years before the COVID-19 diagnosis was not independently associated with COVID-19 outcomes (Figure 1 and Table 2). However, when examining the receipt of PPSV23 only within the last five years, the estimated OR of severe respiratory or critical care outcomes versus non-severe outcomes (Outcome 1), was 1.05 (95% CI: 1.01, 1.08), and the estimated OR for severe respiratory or critical care outcomes versus severe non-respiratory outcomes (Outcome 2), was 1.12 (95% CI: 1.05,1.20). The unexpected direction and magnitude of these results indicated residual bias not adequately accounted for by the model. Similar findings were observed for the receipt of PCV13 on progression to respiratory severe or critical care outcomes from severe non-respiratory and non-severe outcomes (Outcomes 2 and 4) (Figure 1 and Table 2). When looking at the combined effect of PCV13 and PPSV23 within or more than five years from the COVID-19 diagnosis, the results suggest a smaller effect for all outcomes, with a negative direction for Outcome 2 and positive directions for the other outcomes (Table 2).

**Figure 1.**
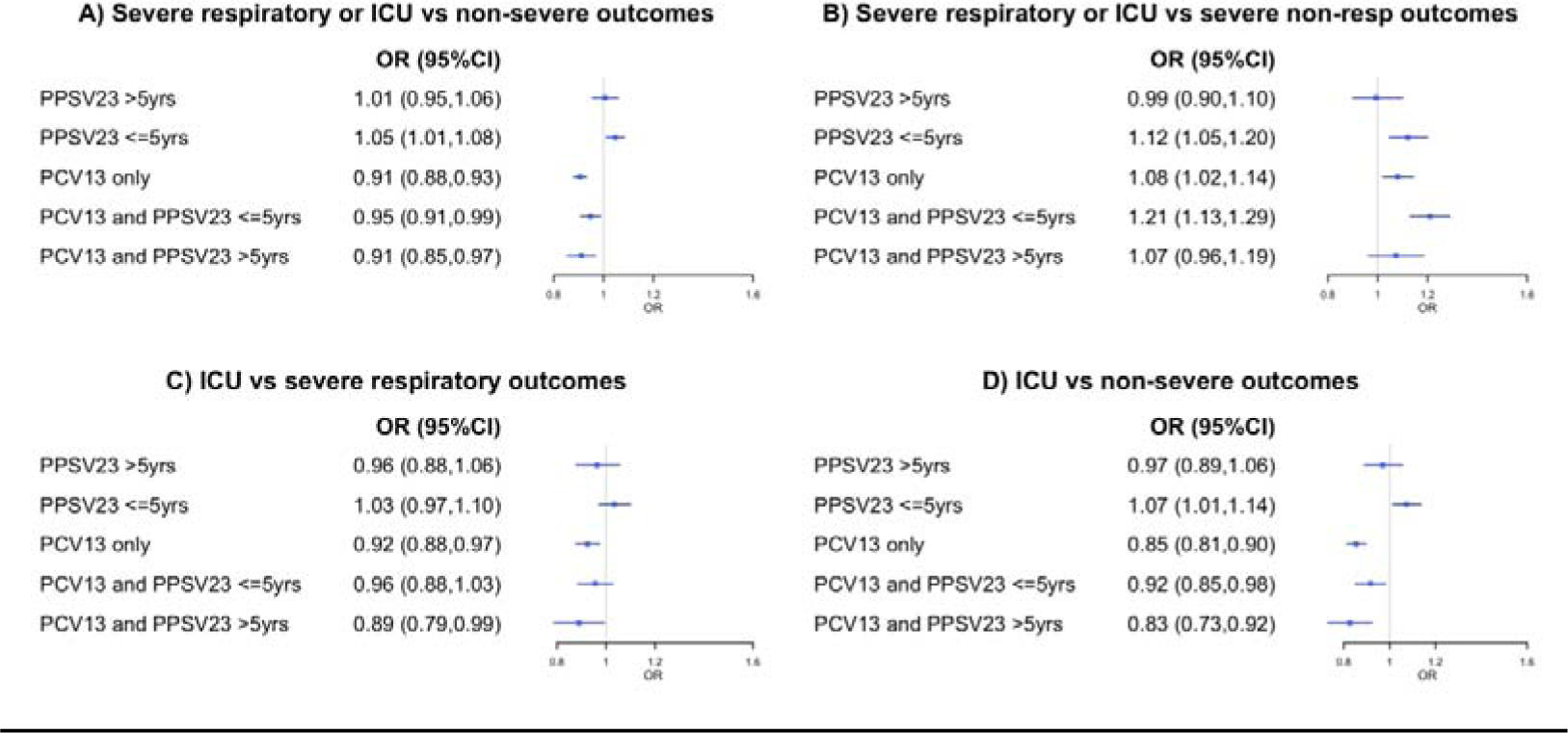
Associations between PCV13 and PPSV23 vaccination and COVID-19 severity. Estimates of the association between any 13-valent pneumococcal vaccine (PCV13) receipt, any 23-valent pneumococcal polysaccharide vaccine (PPSV23) receipt before or after 5 years from the first COVID-19 diagnosis, and receipt of PCV13 and PPSV23 (relative to no pneumococcal vaccination), against progression to severe COVID-19 outcomes. The panels represent four different outcomes: Outcome 1: severe respiratory outcomes or critical care versus non-severe outcomes (panel A). Outcome 2: severe respiratory outcomes or critical care versus severe non-respiratory outcomes (panel B). Outcome 3: critical care versus severe respiratory outcomes (panel C). Outcome 4: critical care versus non-severe outcomes (panel D). Lines (and numbers in parentheses) signify 95% confidence intervals around maximum likelihood estimates (points). Abbreviations: OR, Odds Ratio; CI, confidence interval; PCV13, 13-valent pneumococcal vaccine; PPSV23, 23-valent pneumococcal polysaccharide vaccine.

**Table 2:** Associations between PCV13 and PPSV23 vaccination and COVID-19 severity. Outcome 1: severe respiratory outcomes or critical care versus non-severe outcomes. Outcome 2: severe respiratory outcomes or critical care versus severe non-respiratory outcomes. Outcome 3: critical care versus severe respiratory outcomes Outcome 4: critical care versus non-severe outcomes ** Reference is the unvaccinated category.

| Exposure** | Endpoint (Odds ratio with 95% CI) |  |  |  |
| --- | --- | --- | --- | --- |
|  | Outcome 1 | Outcome 2 | Outcome 3 | Outcome 4 |
| PCV13 only | 0.91<br>(0.88,0.93) | 1.08 (1.02,1.14) | 0.92<br>(0.88,0.97) | 0.85<br>(0.81, 0.90) |
| PPSV23 <= 5years only | 1.05<br>(1.01, 1.08) | 1.12 (1.05,1.20) | 1.03<br>(0.97,1.10) | 1.07<br>(1.01, 1.14) |
| PPSV23 > 5years only | 1.01 (0.95,1.06) | 0.99 (0.90,1.10) | 0.96<br>(0.88, 1.06) | 0.97<br>(0.89, 1.06) |
| PCV13 + PPSV23 <= 5years | 0.95 (0.91, 0.99) | 1.21<br>(1.13, 1.29) | 0.96<br>(0.88, 1.03) | 0.92<br>(0.85, 0.98) |
| PCV13 + PPSV23 > 5years | 0.91 (0.85, 0.97) | 1.07<br>(0.96, 1.19) | 0.89<br>(0.79, 0.99) | 0.83<br>(0.73, 0.92) |
| Alpha variant<br>(original strain is ref) | 1.35 (1.29, 1.40) | 1.27 (1.18,1.37) | 1.39 (1.28,1.51) | 1.71<br>(1.59, 1.85) |
| Delta variant<br>(original strain is ref) | 1.13 (1.08, 1.18) | 1.52 (1.40,1.65) | 1.72 (1.59, 1.88) | 1.66 (1.53, 1.79) |

In our primary analysis, we additionally delineated risk based on COVID-19 variants to investigate whether the association between PCV receipt and COVID-19 severity varied with the original strain, the Alpha variant, and the Delta variant. During both the Alpha and Delta variant phases, there was an elevated risk of all COVID-19 severity outcomes compared to the period when the original strain dominated. The estimated OR of severe respiratory or critical care outcomes versus non-severe outcomes was 1.35 (95% CI: 1.29, 1.40) during the Alpha-variant period and slightly lower during the Delta-variant period (OR: 1.13 with 95% CI: 1.08, 1.18) compared to the original strain period (Table 2). The estimated OR of severe respiratory or critical care outcomes versus non-respiratory severe outcomes was 1.52 during the Delta-variant phase (95% CI: 1.40,1.65) and 1.27 during the Alpha-variant phase (95% CI: 1.18,1.37), relative to the initial COVID-19 period (Table 2). Finally, there was an increase in the odds of critical care versus severe respiratory symptoms (OR=1.39 with 95%CI: 1.28,1.51 and OR=1.72 with 95% CI: 1.59, 1.88, for the Alpha and Delta variants periods respectively), and non-severe symptoms (OR=1.71 with 95%CI: 1.59,1.85 and OR=1.66 with 95% CI: 1.53, 1.79, for the Alpha and Delta variants periods respectively), in comparison to the original strain period (Table 2). In sensitivity analyses, we ran separate models based on the different COVID-19 variants, and the results were consistent with our main analyses (Tables S4, S5, and S6).

To address potential residual confounding, we conducted two sensitivity analyses employing negative-control associations between receipt of influenza vaccine or zoster vaccine and their respective associations with COVID-19 severity, while adjusting for other sources of confounding and excluding PCV13 and PPSV23 from the model (Tables S7 and S8). We observed weak associations between influenza or zoster vaccines and progression to severe respiratory or critical care outcomes from non-severe outcomes (Outcome 1) and between progression to critical care from severe respiratory outcomes and non-severe outcomes (Outcomes 3 and 4) (Tables S7 and S8). The observed associations had similar magnitude to the one observed for the main analysis suggesting that the results are artefactual. Additionally, when examining the association between receipt of influenza or zoster vaccines and odds of severe respiratory or critical care outcomes versus severe non-respiratory outcomes, evidence of residual confounding was apparent (OR: 1.16 with 95%CI: 1.02,1.32 and OR: 1.28 with 95%CI: 1.03,1.62, respectively) (Tables S7 and S8).

## Discussion

In our observational study of US adults aged >= 65 years, we found moderate to no association between the receipt of PCV13 and COVID-19 severity, after adjusting for potential sources of confounding. Similarly, the receipt of PPSV23 vaccine showed no protective association with COVID-19 severity across these outcomes. Our analyses adjusted for comorbid conditions, evidence of previous severe respiratory conditions, and stratification by COVID-19 strain period.

Before starting this study, we had assumed that the most likely mechanism for a link between PCV13 and COVID-19 would be in preventing progression to more severe outcomes. This would occur because vaccination against the bacteria would prevent secondary bacterial infections, as are seen for influenza. However, since the association between PCV13 receipt and progression to more severe outcomes is modest, it seems unlikely that this is the mechanism. It is possible that the initial estimates of the association between PCV13 and COVID-19 has unmeasured residual confounding, despite using a strong study design and extensive checks on the credibility of the estimates [6]. Another possibility would be if PCV13 protects against infection with SARS-CoV-2. A recent study suggests that there could be a synergistic association between pneumococcus and SARS-CoV-2 in the upper airways [18], so potentially preventing colonization with vaccine-targeted serotypes could lower the risk of acquiring SARS-CoV-2. Further longitudinal studies are needed to verify this relationship.

Our findings showing modest effects of PCV13 on specific COVID-19 severity outcomes, especially in the progression from non-severe symptoms to severe respiratory symptoms, necessitates further exploration. The negative control associations that we evaluated suggest that these associations could be an artifact of residual confounding (Tables S7 and S8 in the supplementary file). There was a high level of overlap between individuals receiving influenza vaccine and PCV13 vaccine (Table 1), though incorporating influenza vaccine as a covariate in the model rather than a negative control association did not improve the results. Our results are also consistent with the findings of Lewnard et al. [6], who estimated that PCV13 was effective against COVID-19 infection, hospitalization, and death, but with a similar effect size for each outcome. The similar effect sizes across outcomes described in that study suggest that PCV13 does not affect progression to more severe outcomes, consistent with our findings.

Distinguishing between PPSV23 vaccination within five years and more than five years from COVID-19 diagnosis aimed to account for potential waning of vaccine-induced immunity. However, in both scenarios, we found no negative association between PPSV23 vaccine and COVID-19 severity, in agreement with previous studies that established PPSV23’s limited efficacy in preventing pneumococcal pneumonia.

We adjusted our analyses by COVID-19 strain to elucidate different risks of COVID-19 severity during the Alpha and Delta variant phases compared to the original strain and to remove potential confounding by time. Notably, an elevated risk of progression to more severe outcomes was evident during the Alpha and Delta variant periods across all considered outcomes.

Our study has inherent limitations primarily due to the observational nature of the data. The non-randomized nature of PCV exposure, with vaccine recipients being older and having more comorbidities, introduces potential association bias due to confounding. Although we adjusted for several sources of bias, some estimates still indicate residual confounding, for example, the estimated OR for Outcomes 1 and 2 were positive, suggesting an artifactual negative effect of the PPSV23 vaccine (Table 2). Additionally, since medical claims are financial documents rather than health records, they may not contain complete medical diagnosis information on patients. Further research is required to understand the pneumococcal-coronavirus interactions and the role of PCV vaccination [6].

### Conclusions

In conclusion, our study contributes to understanding the interplay between pneumococcal vaccination and the progression to severe COVID-19 outcomes, indicating a modest to limited association. Future investigations should prioritize identifying vaccines with potential efficacy in enhancing immune response against both heterologous and emerging pathogens.

## Data Availability

All data produced in the present study are available upon reasonable request to the authors

## Ethics declaration

This work was approved by the Institutional Review Board of Georgetown University as Study # STUDY00002324.

## Availability of code and data

The main code can be found in the following github repository: https://github.com/weinbergerlab/PCV-analysis

## Supplementary Appendix

### Further details on the data source

We used Limited Data Set (LDS) files from the Centers for Medicare & Medicaid Services (CMS) from the following files: Inpatient, Outpatient, Skilled Nursing Facility, and Carrier (which contain claims from services provided by professional providers including physicians, physician assistants, clincal social workers and nurse practioners). The Inpatient, Outpatient, and Skilled Nursing Facility files contain 100% of the claims, whereas the Carrier files contain a 5% sample. Combined, the claims files spanned the period from 2014 to 2021. We also had access to the 2014-2016 Denominator LDS files and the 2017-2021 Master Beneficiary Summary File (MBSF) LDS, which provide demographic information on all Medicare beneficiaries.

We formed the study cohort using a set of year-based criteria to guarantee complete, continuous observation of a patient’s medical history (Figure S1). For the years 2014-2016, we included patients with at least 1 claim per year in the 5% Carrier data. For the years 2017-2021, we included patients marked each year with the 5% sample indicator in the MBSF data. We then retained the intersection of these two groups, leaving only patients continuously and completely observed from 2014 to 2021 in the available LDS data (n = 1,046,634). Finally, we reduced this group to those with a COVID-19 diagnosis–designated by the presence of a U07.1 International Classification of Diseases, 10th Revision (ICD-10) code–any time from April 4, 2020 to December 31, 2021 (n = 90,070).

We also used the aforementioned claims data to designate the following characteristics for each member of the cohort: influenza vaccination, pneumococcal conjugate vaccination, zoster vaccination, COVID-19 outcome severity, obesity, and other comorbidities. For all three vaccines, we required the presence of at least one vaccine-specific Current Procedural Terminology (CPT) or Healthcare Common Procedure Coding System (HCPCS) code, provided that the code(s) occurred in a claim before the initial COVID-19 diagnosis (Table S1). COVID-19 outcome severity was split into four mutually exclusive groups of increasing severity: non-severe, non-respiratory severe, respiratory severe, and ICU critical care. Each of these groups required the presence of a U07.1 ICD-10 code along with a unique set of outcome-related codes (Table S2). Additionally, outcome-related diagnosis codes must have co-occurred with a U07.1 diagnosis code and must have fallen within six months of the cohort member’s initial COVID-19 diagnosis. Lastly, we flagged cohort members for obesity and a set of comorbidities based on the presence of specific ICD-10 codes, given that they occurred before the initial COVID-19 diagnosis (Table S3).

**Figure S1.**
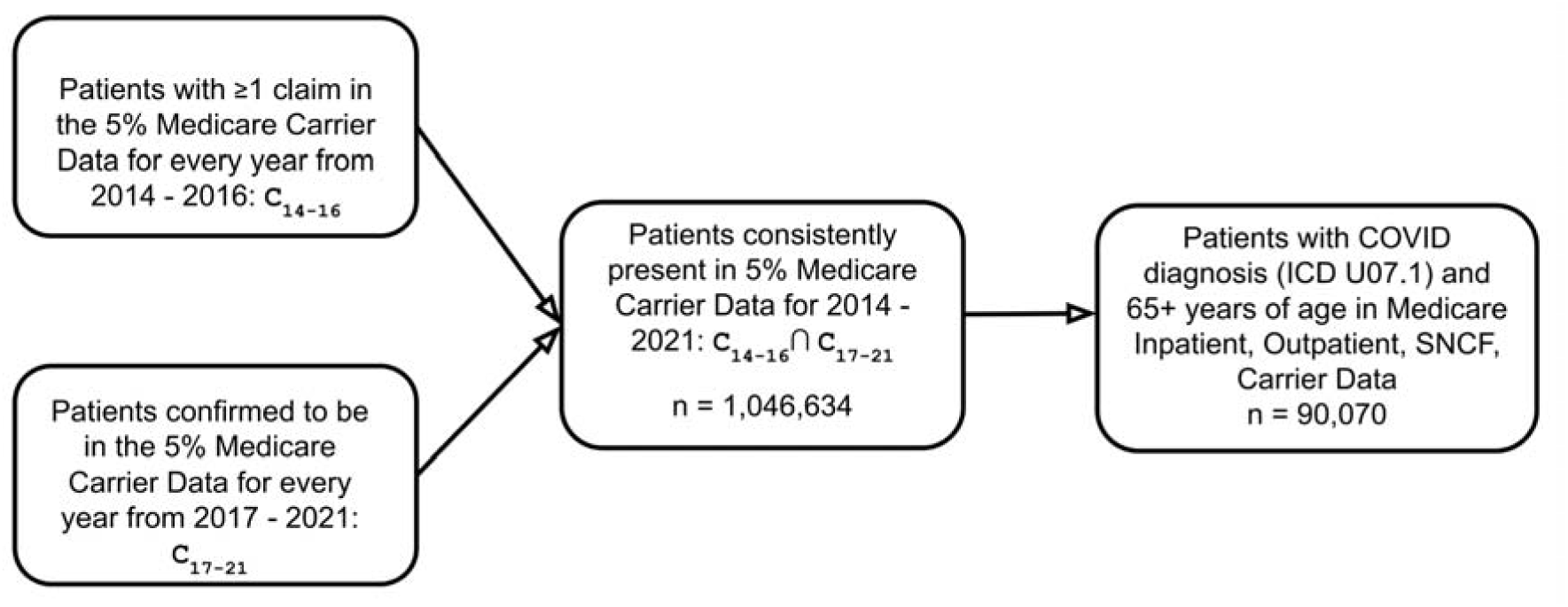
Flowchart describing the construction of the study cohort. We had access to the 100% CMS LDS Inpatient data, 100% CMS LDS Outpatient data, 100% CMS LDS Skilled Nursing Facility data, and the 5% CMS LDS Carrier data sample. To ensure complete observation of patient medical history, we constructed a cohort of continuously observed individuals in the Carrier data (for whom we had complete Inpatient, Outpatient, and Skilled Nursing Facility data). CMS does not provide confirmation about the patients that are in the 5% Carrier data sample for the years 2014 to 2016. Thus, we used the presence of claims in those years as evidence of continuous inclusion in the sample.

**Table S1.** COVID-19 severity definitions. “*” denotes all sub-codes that have the designated root code. Symptom codes may co-occur with the initial COVID-19 diagnosis or occur in claims (also containing U07.1 code) within 6 months of the initial U07.1 code. Patients may have one or more codes to qualify for a given category and are assigned to the highest severity category for which they qualify.

| COVID-19 severity category | ICD-10/CPT code definition |
| --- | --- |
| Non-severe | • U07.1: COVID infection |
| Severe non-respiratory | • U07.1: COVID infection **AND**<br>• At least one of the following:<br>○ A86: unspecified viral encephalitis<br>○ G93.4*: other and unspecified encephalopathy<br>○ I21.*: acute myocardial infarction<br>○ I42.*: cardiomyopathy<br>○ I49.*: cardiac arrhythmias<br>○ I50.*: heart failure<br>○ I63.*: cerebral infarction<br>○ N17.*: acute kidney failure<br>○ R65.2*: severe sepsis<br>○ R74.0*: non-specific elevation of levels of transaminase and lactic acid dehydrogenase |
| Severe Respiratory | • U07.1: COVID infection **AND**<br>• At least one of the following:<br>○ A40.3: sepsis due to *Streptococcus pneumoniae*<br>○ B95.3: *Streptococcus pneumoniae* as the cause of diseases classified elsewhere<br>○ I26.*: pulmonary embolism<br>○ J06.9: acute upper respiratory infection, unspecified<br>○ J12.81: pneumonia due to SARS-associated coronavirus |
|  | ○ J12.82: pneumonia due to coronavirus disease 2019<br>○ J12.89: other viral pneumonia<br>○ J12.9: viral pneumonia, unspecified<br>○ J13: pneumonia due to Streptococcus pneumoniae<br>○ J18.1: lobar pneumonia, unspecified organism<br>○ J18.9: pneumonia, unspecified organism<br>○ J20.8: acute bronchitis due to other specified organisms<br>○ J22: unspecified acute lower respiratory infection<br>○ J40: bronchitis, not specified as acute or chronic<br>○ J80: acute respiratory distress syndrome<br>○ J96.0*: acute respiratory failure<br>○ J98.8: other specified respiratory disorders<br>○ R91.8: other nonspecific abnormal finding of lung field |
| Critical care | ● U07.1: COVID infection **AND**<br>● At least one of the following:<br>○ 99291: critical care service for first 30-74 minutes<br>○ 99292: critical care service for each additional 30 minutes after first 30-74 minutes<br>○ Z99.11: dependence on respirator (ventilator) status |

**Table S2.** Influenza, pneumococcal conjugate, and zoster vaccination CPT/HCPCS code definitions. Vaccination codes must occur in claims from before the initial COVID-19 diagnosis.

| Vaccine | CPT/HCPCS code definition |
| --- | --- |
| Influenza vaccine | 90630, 90653, 90654, 90655, 90656, 90657, 90658, 90660, 90661, 90662, 90664, 90666, 90667, 90668, 90672, 90673, 90674, 90682, 90685, 90686, 90687, 90688, 90689, 90694, 90756, G0008, Q2034, Q2035, Q2036, Q2037, Q2038, Q2039 |
| Pneumococcal conjugate vaccine | 90670, 90732 |
| Zoster vaccine | 90736, 90750 |

**Table S3.** Obesity and comorbidity ICD-10 codes. “*” denotes all sub-codes that have the designated root code. We ensure that claims containing these codes occur before the patient’s first COVID-19 diagnosis.

| Condition | ICD-10 code definition |
| --- | --- |
| Obesity | ● At least one of the following:<br>○ Z68.3*: BMI 30-39, adult<br>○ Z68.4*: BMI 40 or greater, adult |
| Comorbidities | ● At least one of the following: |
|  | ○ B20: Human immunodeficiency virus (HIV) disease<br>○ D84.9: immunodeficiency, unspecified<br>○ E10.*: Type 1 diabetes mellitus<br>○ E11.*: Type 2 diabetes mellitus<br>○ E84.*: cystic fibrosis<br>○ F03.*: unspecified dementia<br>○ I25.1*: atherosclerotic heart<br>○ I26.*: pulmonary embolism<br>○ I27.0: primary pulmonary hypertension<br>○ I42.*: cardiomyopathy<br>○ I50.*: heart failure<br>○ disease of native coronary artery<br>○ J44.*: other chronic obstructive pulmonary disease<br>○ J47.*: bronchiectasis<br>○ J84.*: other interstitial pulmonary diseases<br>○ K70.0: alcoholic fatty liver<br>○ K74.*: fibrosis and cirrhosis of liver<br>○ K75.4: autoimmune hepatitis<br>○ K76.0: fatty (change of) liver, not elsewhere classified<br>○ N18.*: chronic kidney disease (CKD)<br>○ Z87.891: personal history of nicotine dependence<br>○ Z94.*: transplanted organ and tissue status |
| Past severe respiratory illness | ● At least one of the following before the earliest COVID-19 diagnosis:<br>○ A40.3: sepsis due to *Streptococcus pneumoniae*<br>○ B95.3: *Streptococcus pneumoniae* as the cause of diseases classified elsewhere<br>○ I26.*: pulmonary embolism<br>○ J06.9: acute upper respiratory infection, unspecified<br>○ J12.81: pneumonia due to SARS-associated coronavirus<br>○ J12.82: pneumonia due to coronavirus disease 2019<br>○ J12.89: other viral pneumonia<br>○ J12.9: viral pneumonia, unspecified<br>○ J13: pneumonia due to *Streptococcus pneumoniae*<br>○ J18.1: lobar pneumonia, unspecified organism<br>○ J18.9: pneumonia, unspecified organism<br>○ J20.8: acute bronchitis due to other specified organisms<br>○ J22: unspecified acute lower respiratory infection<br>○ J40: bronchitis, not specified as acute or chronic<br>○ J80: acute respiratory distress syndrome<br>○ J96.0*: acute respiratory failure<br>○ J98.8: other specified respiratory disorders<br>○ R91.8: other nonspecific abnormal finding of lung field |

**Table S4:** Three different models showing associations between 1) PCV13 and PPSV23 vaccination and COVID-19 severity; 2) influenza vaccine and COVID-19 severity; 3) zoster vaccine and COVID-19 severity during the original COVID-19 strain period. Outcome 1: severe respiratory outcomes or critical care versus non-severe outcomes. Outcome 2: severe respiratory outcomes or critical care versus severe non-respiratory outcomes. Outcome 3: critical care versus severe respiratory outcomes. Outcome 4: critical care versus non-severe outcomes. ** Reference is the unvaccinated category. N=14,856

| Exposure** | Endpoint (Odds ratio with 95% CI) |  |  |  |
| --- | --- | --- | --- | --- |
|  | Outcome 1 | Outcome 2 | Outcome 3 | Outcome 4 |
| 1) <u>Primary model</u> |  |  |  |  |
| PCV13 only | 0.95<br>(0.88,1.02) | 1.16 (1.02,1.32) | 0.79<br>(0.68,0.92) | 0.80 (0.69, 0.92) |
| PPSV23 <= 5years only | 1.11<br>(1.02, 1.21) | 1.14 (0.98,1.35) | 1.07<br>(0.90, 1.27) | 1.19 (1.01, 1.40) |
| PPSV23 > 5years only | 1.11 (0.96,1.28) | 1.42 (1.07,1.91) | 1.02<br>(0.76, 1.35) | 1.15 (0.87, 1.49) |
| PCV13 + PPSV23 <= 5years | 1.06 (0.95,1.16) | 1.33 (1.13, 1.52) | 0.85<br>(0.64, 1.05) | 0.95 (0.75,1.14) |
| PCV13 + PPSV23 > 5years | 1.05<br>(0.89, 1.21) | 1.64<br>(1.33, 1.96) | 1.12<br>(0.50, 0.81) | 0.91<br>(0.62, 1.21) |
| 2) <u>Negative control analysis with flu vaccination</u> |  |  |  |  |
| Flu vaccine | 1.06<br>(0.99,1.15) | 1.15 (1.01,1.32) | 0.86<br>(0.74,0.99) | 0.95<br>(0.82, 1.09) |
| 3) <u>Negative control analysis with zoster vaccination</u> |  |  |  |  |
| Zoster vaccine | 1.00 (0.78,1.28) | 1.41 (0.87,2.42) | 0.52<br>(0.27,0.91) | 0.64 (0.34, 1.09) |

**Table S5:** Three different models showing associations between 1) PCV13 and PPSV23 vaccination and COVID-19 severity; 2) influenza vaccine and COVID-19 severity; 3) zoster vaccine and COVID-19 severity during the COVID-19 Alpha strain period. Outcome 1: severe respiratory outcomes or critical care versus non-severe outcomes. Outcome 2: severe respiratory outcomes or critical care versus severe non-respiratory outcomes. Outcome 3: critical care versus severe respiratory outcomes. Outcome 4: critical care versus non-severe outcomes. ** Reference is the unvaccinated category. N=43,547

| Exposure** | Endpoint (Odds ratio with 95% CI) |  |  |  |
| --- | --- | --- | --- | --- |
|  | Outcome 1 | Outcome 2 | Outcome 3 | Outcome 4 |
| 4) <u>Primary model</u> |  |  |  |  |
| PCV13 only | 0.97<br>(0.93,1.01) | 1.15 (1.06,1.24) | 0.92<br>(0.86,0.99) | 0.92 (0.86, 0.99) |
| PPSV23 <= 5years only | 1.06<br>(1.01, 1.12) | 1.15 (1.04,1.26) | 1.03<br>(0.95, 1.12) | 1.09 (1.01, 1.18) |
| PPSV23 > 5years only | 1.05 (0.98,1.14) | 0.99 (0.86,1.13) | 0.99<br>(0.86, 1.12) | 1.02 (0.90, 1.16) |
| PCV13 + PPSV23 <= 5years | 1.03 (0.97, 1.09) | 1.32<br>(1.21,1.43) | 0.95<br>(0.85, 1.05) | 1.00 (0.91, 1.10) |
| PCV13 + PPSV23 > 5years | 1.02 (0.94, 1.10) | 1.13 (0.97,1.29) | 0.91 (0.76,1.06) | 0.94 (0.80, 1.08) |
| 5) <u>Negative control analysis with flu vaccination</u> |  |  |  |  |
| Flu vaccine | 1.00<br>(0.96,1.04) | 1.18 (1.09,1.27) | 0.93<br>(0.86,1.00) | 0.96 (0.89, 1.03) |
| 6) <u>Negative control analysis with zoster vaccination</u> |  |  |  |  |
| Zoster vaccine | 0.85 (0.73,0.98) | 1.31 (0.96,1.83) | 0.83<br>(0.63,1.09) | 0.72 (0.55,0.93) |

**Table S6:** Three different models showing associations between 1) PCV13 and PPSV23 vaccination and COVID-19 severity; 2) influenza vaccine and COVID-19 severity; 3) zoster vaccine and COVID-19 severity during the COVID-19 Delta strain period. Outcome 1: severe respiratory outcomes or critical care versus non-severe outcomes. Outcome 2: severe respiratory outcomes or critical care versus severe non-respiratory outcomes. Outcome 3: critical care versus severe respiratory outcomes. Outcome 4: critical care versus non-severe outcomes. ** Reference is the unvaccinated category. N=31,667

|  | Endpoint (Odds ratio with 95% CI) |
| --- | --- |

| Exposure** | Outcome 1 | Outcome 2 | Outcome 3 | Outcome 4 |
| --- | --- | --- | --- | --- |
| 7) <u>Primary model</u> |  |  |  |  |
| PCV13 only | 0.80<br>(0.76,0.85) | 0.93 (0.83,1.03) | 0.97<br>(0.89,1.07) | 0.79 (0.73, 0.85) |
| PPSV23 <= 5years only | 1.00<br>(0.94, 1.06) | 1.06 (0.93,1.20) | 1.02<br>(0.92, 1.14) | 1.02 (0.93,1.13) |
| PPSV23 > 5years only | 0.93 (0.85,1.01) | 0.88 (0.74,1.05) | 0.92<br>(0.79, 1.06) | 0.89 (0.77,1.02) |
| PCV13 + PPSV23 <= 5years | 0.80 (0.74,0.87) | 0.98 (0.83,1.13) | 1.00 (0.88, 1.12) | 0.81 (0.70, 0.91) |
| PCV13 + PPSV23 > 5years | 0.75 (0.65,0.84) | 0.82 (0.62,1.01) | 0.89 (0.73,1.06) | 0.70 (0.55,0.85) |
| 8) <u>Negative control analysis with flu vaccination</u> |  |  |  |  |
| Flu vaccine | 0.78<br>(0.75,0.82) | 0.84 (0.75,0.93) | 0.95<br>(0.87,1.03) | 0.76 (0.70,0.82) |
| 9) <u>Negative control analysis with zoster vaccination</u> |  |  |  |  |
| Zoster vaccine | 0.85 (0.71,1.01) | 1.20 (0.80,1.88) | 0.95<br>(0.68,1.31) | 0.84 (0.62,1.11) |

**Table S7:** Associations between influenza vaccination and COVID-19 severity. Outcome 1: severe respiratory outcomes or critical care versus non-severe outcomes. Outcome 2: severe respiratory outcomes or critical care versus severe non-respiratory outcomes. Outcome 3: critical care versus severe respiratory outcomes Outcome 4: critical care versus non-severe outcomes ** Reference is the unvaccinated category.

| Exposure** | Endpoint (Odds ratio with 95% CI) |  |  |  |
| --- | --- | --- | --- | --- |
|  | Outcome 1 | Outcome 2 | Outcome 3 | Outcome 4 |

| <u>Flu vaccination (False is the ref, i.e. no record of flu vaccination)</u> |  |  |  |  |
| --- | --- | --- | --- | --- |
| Flu vax | 0.94<br>(0.91,0.96) | 1.05<br>(0.99,1.11) | 0.91<br>(0.87,0.97) | 0.88<br>(0.83, 0.92) |

**Table S8:** Associations between zoster vaccination and COVID-19 severity. Outcome 1: severe respiratory outcomes or critical care versus non-severe outcomes. Outcome 2: severe respiratory outcomes or critical care versus severe non-respiratory outcomes. Outcome 3: critical care versus severe respiratory outcomes Outcome 4: critical care versus non-severe outcomes ** Reference is the unvaccinated category.

| **Exposure\*\*** | **Endpoint (Odds ratio with 95% CI)** |  |  |  |
| --- | --- | --- | --- | --- |
|  | Outcome 1 | Outcome 2 | Outcome 3 | Outcome 4 |
| <u>Zoster vaccination (False is the ref, i.e. no record of flu vaccination)</u> |  |  |  |  |
| Zoster vax | 0.88<br>(0.79,0.97) | 1.28<br>(1.03,1.62) | 0.82<br>(0.67,1.00) | 0.76<br>(0.63, 0.91) |

